# Immune cell distributions in the blood of healthy individuals at high genetic risk of Parkinson’s disease

**DOI:** 10.1101/2024.10.31.24316489

**Authors:** Laura Deecke, David Goldeck, Olena Ohlei, Jan Homann, Ilja Demuth, Lars Bertram, Graham Pawelec, Christina M. Lill

**Author notes:** Corresponding Author:* Christina M. Lill, MD, MSc, Translational Epidemiology Group, Institute of Epidemiology and Social Medicine, University of Münster, Albert-Schweitzer-Campus 1, 48149 Münster, Germany.

## Abstract

The immune system likely plays a key role in Parkinson’s disease (PD) pathophysiology. Thus, we investigated whether immune cell compositions are already altered in healthy individuals at high genetic risk for PD. We quantified 92 immune cell subtypes in the blood of 442 individuals using multicolor flow cytometry. Polygenic risk scores (PGS) for PD were calculated based on genome-wide significant SNPs (n = 87) from a large genome-wide association study (n = 1,530,403). Linear regression analyses did not reveal significant associations between PGS and any immune cell subtype (FDR = 0.05). Nominally significant associations were observed for NKG2C+ B cells (p = 0.026) in the overall sample. Older participants at increased genetic PD risk also showed a higher proportion of myeloid dendritic cells (p = 0.019) and CD27+CD4+ memory T cells (p = 0.043). Several immune cells were nominally statistically associated in women only. These findings suggest that major alterations of immune cells only occur later in the progression of PD.

## Introduction

Idiopathic Parkinson’s disease (PD) is a genetically complex disease determined by a combination and interaction of genetic, environmental and lifestyle factors. The immune system likely plays a critical role in its development (1).Along this line, there is increasing evidence for the involvement of different immune cell subtypes in the pathophysiology of PD (2,3). For example, monocytes are reportedly elevated in PD patients compared to controls, and alterations have also been described for various subsets of T cells (2–4). However, such studies often yield inconsistent results, possibly due to differences in treatment, disease duration, and comorbidities across participants. These discrepancies make it difficult to determine cause-effect relationships, and reverse causation may also be present. Thus, to investigate the earliest changes of immune cell alterations in the course of PD, we explored whether healthy individuals at high genetic risk for PD already show altered immune cell profiles. To this end, we calculated polygenic risk scores (PGS) for PD and analyzed their association with more than 90 immune cell subtypes in the blood of nearly 450 healthy participants from the Berlin Aging Study II (BASE-II; **Figure 1**).

**Figure 1.**
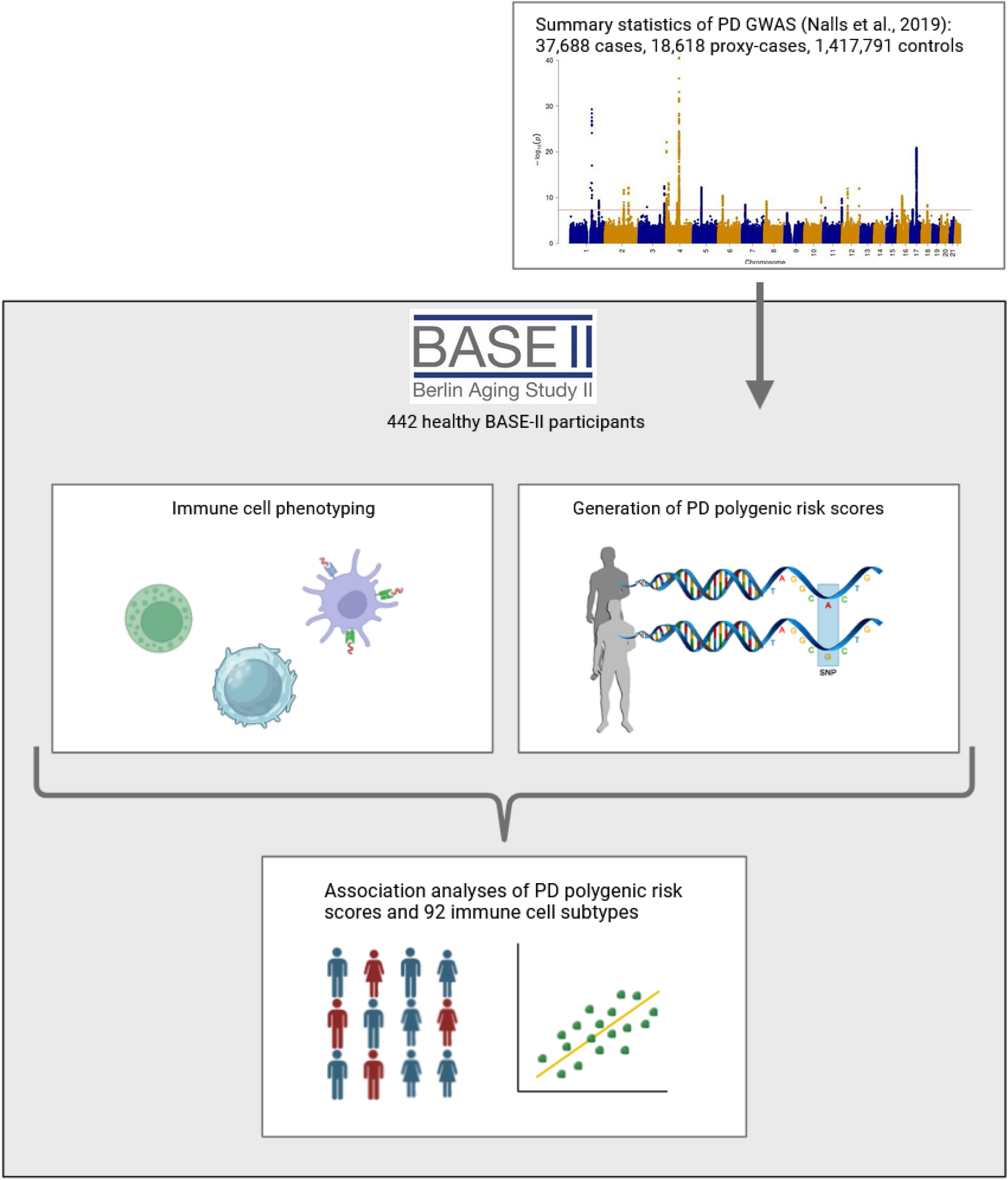
Study overview. Schematic overview of the present study generated using BioRender.com. The Manhattan plot of the genome-wide association study was generated based on the summary statistics made available by Nalls et al. (8).

## Methods

### Study participants

This study included healthy adults from BASE-II, a multi-institutional longitudinal study on characteristics of aging (5). Participants were of European descent, recruited from the Berlin area, and were immunologically (no fever or immune system-related diseases) and cognitively (mini mental state examination test ≥27) healthy with no PD symptoms. Written informed consent was obtained from all participants, and the study was approved by the institutional review boards of all participating institutions. The cohort (n = 442) was composed of a group of older individuals and of younger individuals (**Supplementary Figure 1**) for whom quality-controlled (QC) genetic as well as immune cell data were available.

### Generation and quality control of immune cell data by flow cytometry

The isolation of peripheral mononuclear blood cells (PBMC) from whole blood of BASE-II participants and flow cytometry using two different panels was performed as previously described (6,7): In short, ‘panel 1’ mainly contained T cell subtypes, whereas ‘panel 2’ comprised a range of immune cell subtypes including natural killer cells, natural killer T cells, monocytes, myeloid-derived suppressor cells (MDSC), B cells, as well as general T cell populations (**Supplementary Table 1-2**). Antibodies used for the isolation of individual cell types in the panels can be found in **Supplementary Table 2**. Cells were acquired with a 3 laser BD LSRII (BD biosciences) flow cytometer and DIVA6 software. Data were analyzed with FlowJo version 7.5 (TreeStar) (6,7). The immune cells were quantified as blood proportions. Where applicable, the proportions of immune cells were transformed using log10, root, log(100-x) and square transformations, respectively, and z-transformed (**Supplementary Tables 1-2**).

### Processing of genome-wide SNP data and generation of PGS

Genome-wide SNP data were generated using the Affymetrix Array 6.0 and subjected to standard QC as described elsewhere (7). Ungenotyped genotypes were imputed using the Haplotype Reference Consortium reference (7). This resulted in 7,512,709 SNPs with a minor allele frequency (MAF) of ≥0.01 and an imputation r^2^ ≥0.3. We calculated the PD PGS (**Supplementary Figure 1)** based on 87 of the 90 genome-wide significantly and independently associated SNPs reported in the largest GWAS on PD risk in Caucasian populations (Table S2 from the original study (8)). To maximize the number of PD risk SNPs included, we relaxed our standard QC thresholds for some variants: This related to two variants with MAF <0.01 (i.e., rs114138760 and rs76763715; rs34637584 was monomorphic in our study), and two SNPs showing an imputation r^2^ slightly below 0.3 (r^2^_rs62053943_ = 0.25, r^2^_rs117615688_ = 0.16). Only two SNPs in the human leukocyte antigen (HLA) region were eventually excluded (imputation r^2^ <0.013). Of note, the 90 independent SNPs described in the original study (8) had been identified using the conditional and joint analysis method (COJO) (9) and included a few SNPs located in the same locus. Thus, in sensitivity analyses, we calculated alternate PGS by i) only including the most significantly associated SNP per locus (±1Mb; n = 75 SNPs) and ii) by excluding the four SNPs mentioned above not meeting our stringent QC criteria (**Supplementary Tables 3-4**).

### Statistical analyses

Linear regression analyses of the immune cell distributions on the z-transformed PGS was performed using the lm function in R (10). Analyses were adjusted for sex, age group, and the first four genetic principal components (derived from principal component analyses) to adjust for subtle differences in ancestry. The number of principal components was determined using scree plots. Subgroup analyses were performed for older participants only and stratified by sex. Sensitivity analyses included adjustments for cytomegalovirus (CMV) status and modifications of the PGS calculation (see above). The false discovery rate (FDR) was controlled at 5%.

## Results

The BASE-II dataset analyzed here (n = 442, 59% women) consisted of 305 older individuals (59% women, median age: 69 years, range: 60-82 years) and 137 younger individuals (59% women, median age: 29 range, range: 23-35 years, **Supplementary Figure 1**).

Linear regression analyses in the BASE-II participants did not reveal any significant associations between the PGS and the 92 immune cell subtypes after FDR control (FDR = 5%), neither in the overall (n=442) nor in the subgroup analyses, i.e., stratifying by sex (i.e., 262 women and 180 men) or analyzing only the older age group (n=305; **Supplementary Table 4**). However, we observed several nominally significant associations: In the overall sample, NKG2C+ B cells tended to be increased in individuals at higher genetic PD risk (variance explained [ΔR2] = 1.5%, p = 0.026) with consistent effect directions across subgroups and the strongest effect observed in women (ΔR2 = 2.9%, p = 0.014, **Table 1**). Furthermore, when restricting our analyses to participants from the old age group, i.e., those closer to a potential onset of PD, higher genetic PD risk was associated with increased proportions of myeloid dendritic cells (ΔR2 = 2.5%, p = 0.019) and CD27+ CD4+ memory T cells (ΔR2 = 1.6%, p = 0.043). In addition, women showed inverse associations between PD risk and two immune cell subtypes (myeloid-derived suppressor cells type I [MDSC1], ΔR2 = 2.4%, p = 0.024; lineage-negative HLA-DR-cells [Lin-HLA-DR-], ΔR2 = 2.0%, p = 0.038), with null effects in men. (**Table 1**). Overall, the variance explained for these nominally significant associations ranged from 1.5% to 2.9%. (**Table 1**). Sensitivity analyses adjusting for CMV status or using the two alternate PGS did not change the results substantially (**Supplementary Table 4**).

**Table 1.**
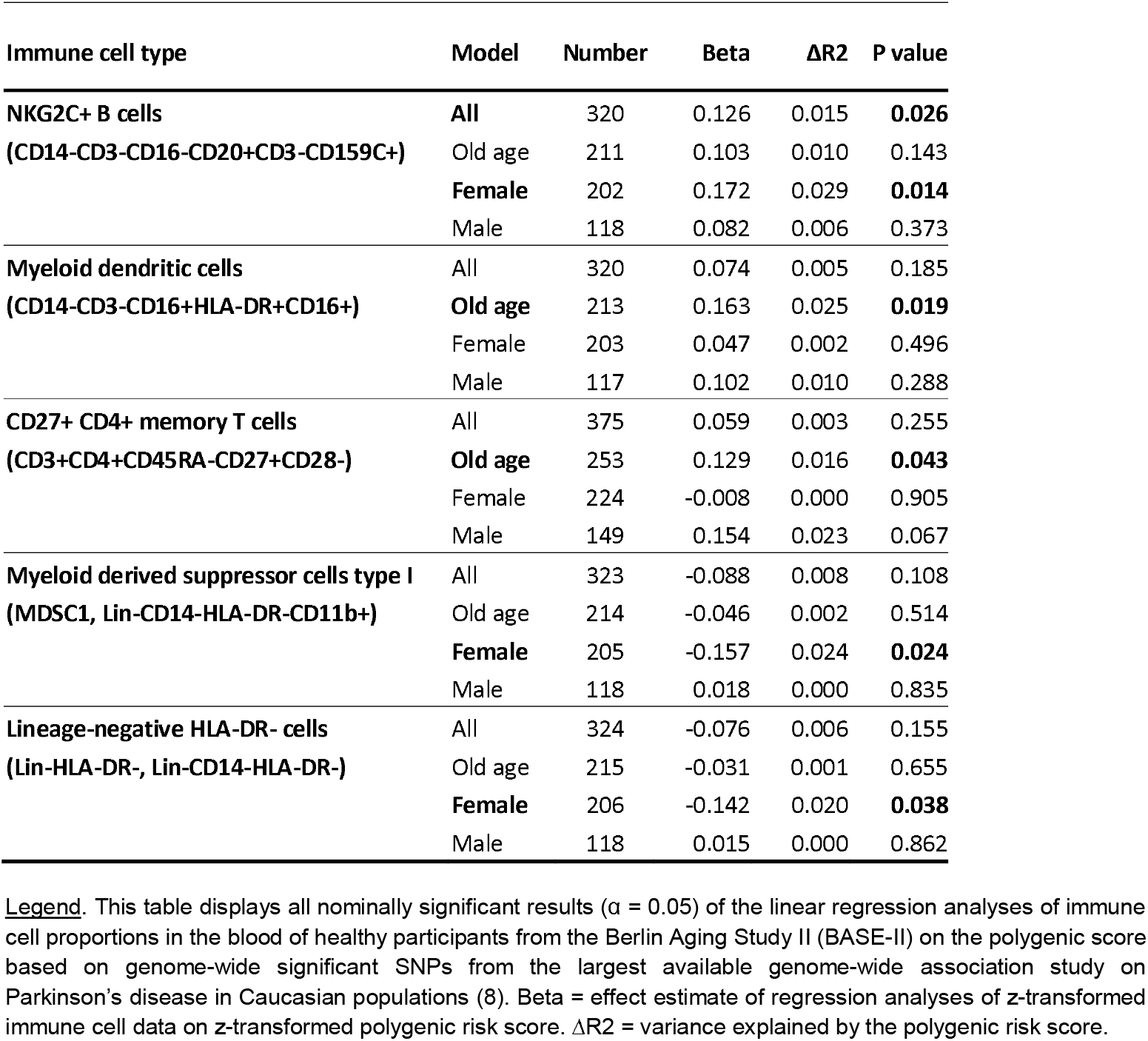
Association of Parkinson’s disease polygenic risk score with immune cell distributions in the blood of healthy BASE-II participants.

## Discussion

To the best of our knowledge, this is the first study to investigate immune cell alterations in healthy individuals at high genetic risk for PD. While no statistically significant changes were detected after FDR control, we observed several nominally significant and functionally relevant alterations, such as increased NKG2C+ B cells (full sample) and higher proportions of myeloid dendritic cells and CD27+ CD4+ memory T cells (old age group). In agreement with the latter observation, CD4+ memory T cells have been reported to be increased in prevalent PD patients compared to controls in previous work (2,11). In contrast, our observation of increased NKG2C+ B cells with PD risk is novel and warrants further investigation. Notably, B cell subtypes other than NKG2C cells have been reported as either increased (12,13) or decreased (2,14) in PD, though findings across studies remain somewhat inconsistent. Myeloid dendritic cells, which present α-synuclein to T cells and trigger adaptive immune responses in PD, showed a tendency for an increase with PD risk in our study but have also been reported to show a decline with respect to disease severity in diagnosed PD patients (15), indicating a complex role in PD. Interestingly, several immune cell subtypes showed stronger alterations in women than men (NKG2C+ B cells) or were altered only in women but not men (MDSC1, Lin-HLA-DR-), possibly hinting at sex-specific compensatory immune mechanisms, although this remains speculative at this point in time.

The main strengths of our study are i) deep standardized immune cell phenotyping, ii) comprehensive clinical characterization of the BASE-II participants, iii) a comparatively large sample size. However, despite being able to analyze nearly 450 individuals, power was limited to detect small differences in immune cell composition. In this context and in at least partial agreement with the existing literature, it is possible that the trends of immune cell alterations observed in our study represent genuine findings. Secondly, we investigated individuals at high genetic risk of developing PD, i.e., we did not consider the impact of lifestyle and environmental PD risk factors and only a fraction of participants may eventually develop PD. The latter may have ‘diluted’ potential prodromal alterations of immune cells in PD pathophysiology. Lastly, we only included participants of European descent, limiting generalizability to other ethnic groups.

In conclusion, immune cell alterations do not seem to be predominant in clinically unaffected participants at high genetic risk of PD. However, our suggestive, nominally significant findings indicating increases of B cell subtypes, myeloid dendritic cells, and memory T cells in individuals at high genetic risk for PD as well as the potential sex-specific effects warrant future validation.

## Author Contributions

Conceptualization: C.M.L.; methodology: L.D., C.M.L.; formal analysis or interpretation of the data: L.D., C.M.L.; data acquisition: D.G., G.P.; writing—original draft preparation: L.D., C.M.L.; writing—review and editing: D.G., O.O., J.H., I.D., L.B., G.P.; supervision, C.M.L. All authors have read and agreed to the published version of the manuscript.

## Data Availability

All summary statistics have been made available in the supplementary material of this manuscript. Subject-level data can be obtained by qualified investigators upon request to the authors.

## Acknowledgements

We are grateful to all BASE-II participants.

## Ethical approval

All BASE-II participants provided written informed consent before participation and the study was conducted in accordance with the Declaration of Helsinki and approved by the Ethics Committee of the Charité-Universitätsmedizin Berlin – approval number EA2/029/09.

