## Supplementary figures and images for "Immune cell distributions in the blood of healthy individuals at high genetic risk of Parkinson’s disease"

### Supplementary Figure 1

1a. Age distribution in the Berlin Aging Study II

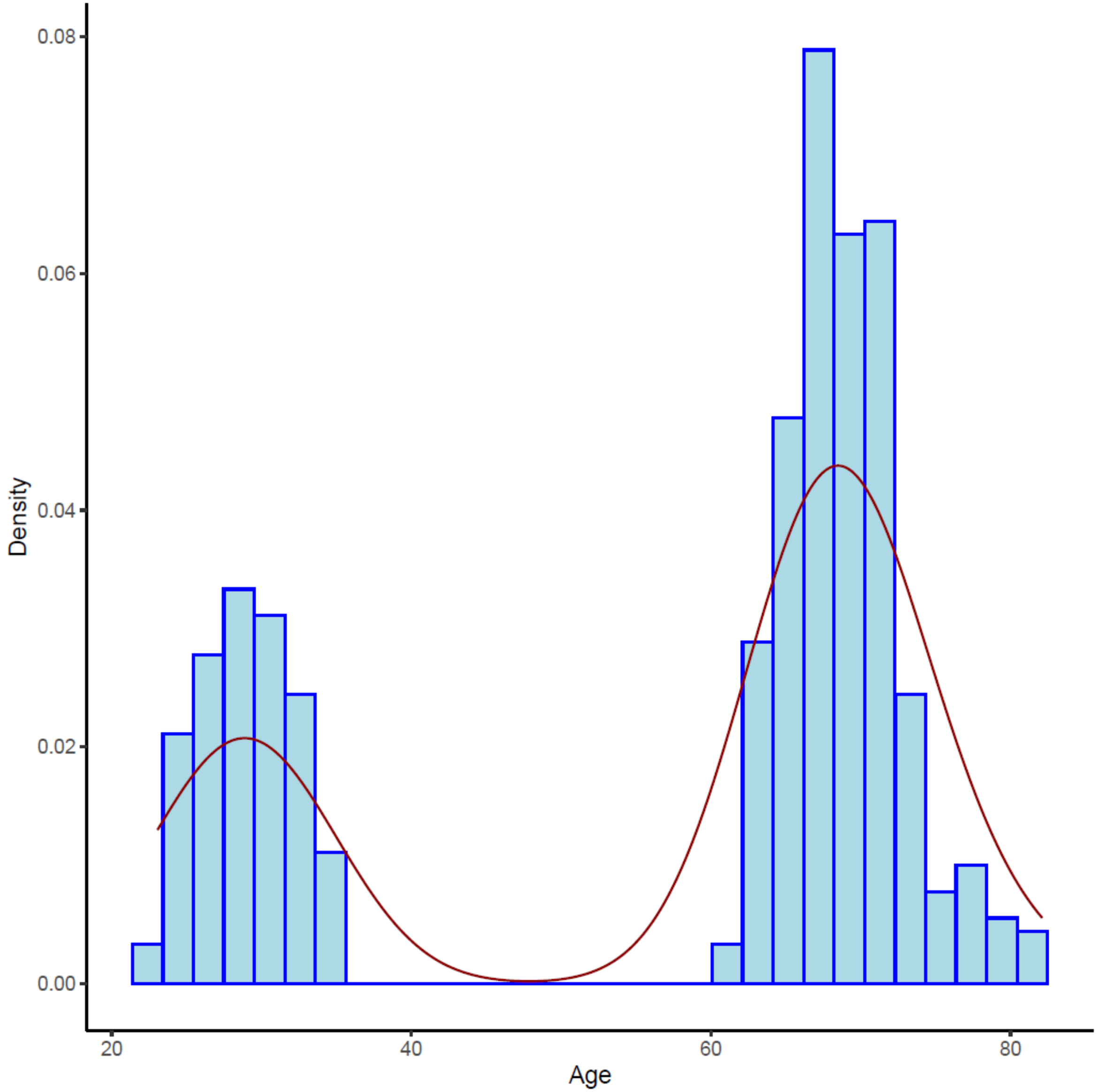


1b. Distribution of the polygenic risk score

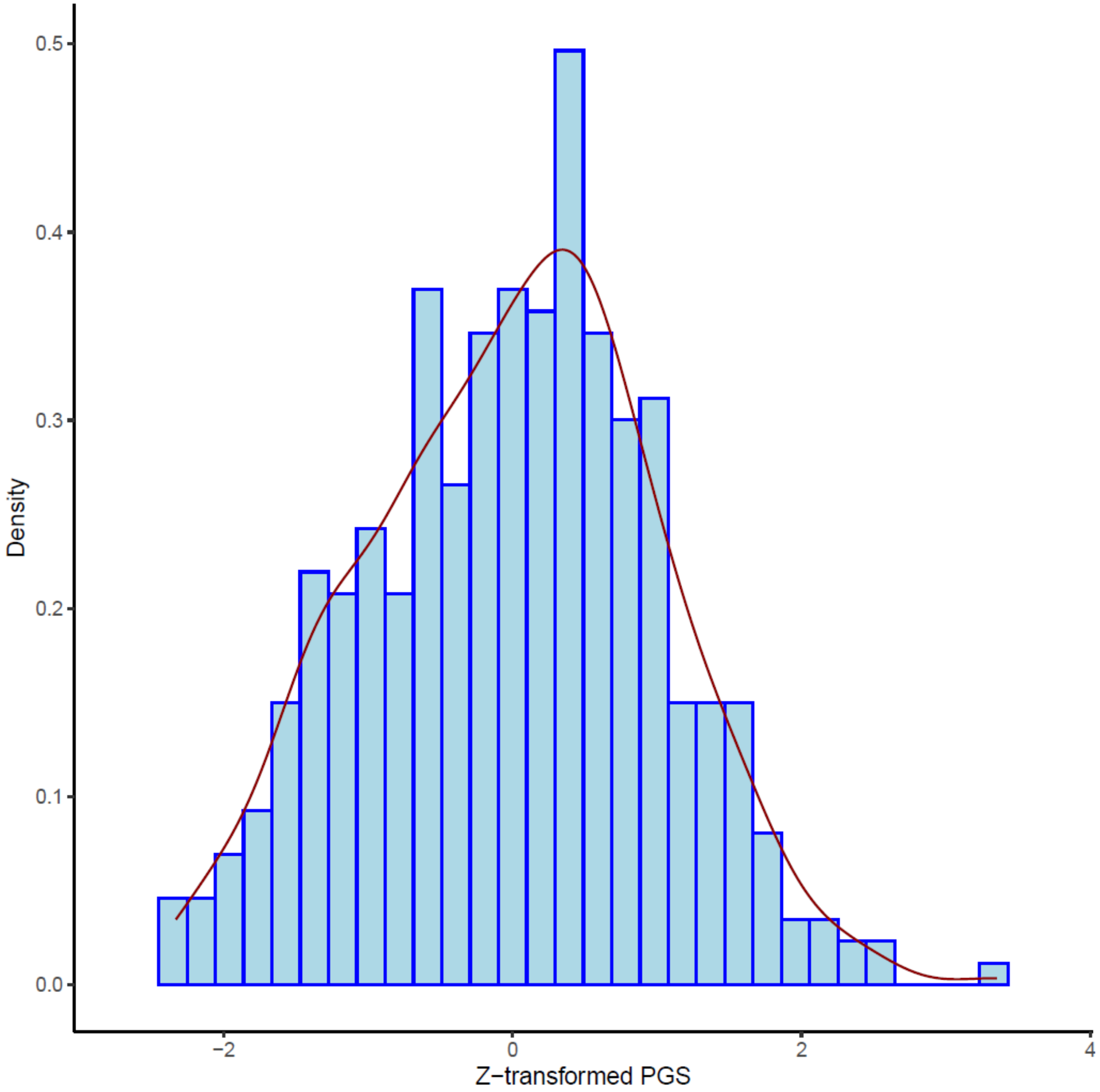
